# Population-level management of Type 1 diabetes via continuous glucose monitoring and algorithm-enabled patient prioritization: Precision health meets population health

**DOI:** 10.1101/2021.05.04.21256647

**Authors:** Johannes O. Ferstad, Jacqueline J. Vallon, Daniel Jun, Angela Gu, Anastasiya Vitko, Dianelys P. Morales, Jeannine Leverenz, Ming Yeh Lee, Brianna Leverenz, Christos Vasilakis, Esli Osmanlliu, Priya Prahalad, David M Maahs, Ramesh Johari, David Scheinker

## Abstract

**Objective:** To develop and scale algorithm-enabled patient prioritization to improve population-level management of type 1 diabetes (T1D) in a pediatric clinic with fixed resources, using telemedicine and remote monitoring of patients via continuous glucose monitor (CGM) data review.

**Research Design and Methods:** We adapted consensus glucose targets for T1D patients using CGM to identify interpretable clinical criteria to prioritize patients for weekly provider review. The criteria were constructed to manage the number of patients reviewed weekly and identify patients who most needed provider contact. We developed an interactive dashboard to display CGM data relevant for the patients prioritized for review.

**Results:** The introduction of the new criteria and interactive dashboard was associated with a 60% reduction in the mean time spent by diabetes team members who remotely and asynchronously reviewed patient data and contacted patients, from 3.2±0.20 to 1.3±0.24 minutes per patient per week. Given fixed resources for review, this corresponded to an estimated 147% increase in weekly clinic capacity. Patients who qualified for and received remote review (n=58) have associated 8.8 percentage points (pp) (95% CI = 0.6–16.9pp) greater time-in-range (70-180 mg/dL) glucoses compared to 25 control patients who did not qualify at twelve months after T1D onset.

**Conclusions:** An algorithm-enabled prioritization of T1D patients with CGM for asynchronous remote review reduced provider time spent per patient and was associated with improved time-in-range.

---

Continuous glucose monitoring (CGM) and data-driven care tools improve patient outcomes.^1–3^ CGM use is recommended for all patients with T1D and augments the capabilities of telemedicine in diabetes care.^4^ Unfortunately, a majority of patients with T1D still self-monitor their blood glucose through finger-sticks. A majority also fail to meet HbA1c targets.^5^ Gaps in diabetes technology use and patient outcomes are further exacerbated by structural racial and socioeconomic disparities.^6,7^

Clinics seeking to expand access to their telemedicine program, especially to underserved populations geographically separated from pediatric endocrinologists, stand to benefit from efficiencies that allow them to serve more patients with their current resources. CGM-enabled telemedicine (synchronous and asynchronous) could expand specialized T1D care to underserved areas through comprehensive care networks.^8,9^ The automation of time-consuming tasks that do not require human expertise can free provider time, thus enabling clinics to care for more patients while improving patient outcomes, without requiring additional resources. Clinic efficiency may improve with algorithms that help providers accurately identify patients with suboptimal glucose control without having to manually review data for the whole clinic population.

In order to facilitate efficient care delivery for patients using CGM, clinics need help analyzing large amounts of CGM data and limiting the total review time required of providers.^10^ Data science and algorithm design have the potential to direct attention to the patients most likely to benefit from provider contact and to fill existing technical deficiencies to enable the Digital Diabetes Clinic.^11,12^ Many tools exist to analyze CGM data for an individual patient.^13–15^ But, to our knowledge, no open-source, CGM manufacturer-independent tools are available to help clinics analyze CGM data and manage their entire T1D clinic population.

In a clinic providing T1D care through synchronous and asynchronous telemedicine based on the review of CGM data, we demonstrate an approach to reduce provider review time per patient by prioritizing patients based on their likelihood of benefiting from an intervention by the care team. We do so by evaluating the impact of an interactive patient CGM glucose data dashboard that makes it possible to expand periodic asynchronous remote CGM review to larger patient populations without increasing provider screen time or negatively affecting patients’ glucose control. We developed an algorithm that ranks patients by their number of flags (i.e., indicators of glucose outside of targets), and then by ascending time in range (TIR). Based on earlier work providing population-level care through the analysis of CGM data, we hypothesized such an algorithm would yield both high sensitivity and high specificity.^1^ The algorithm is based on consensus guidelines, is available through free open-source software, is compatible with data from any manufacturer that support raw data extracts, and allows clinics to specify their own patient review criteria based on their capacity constraints.^16,17^ After deploying the algorithm to identify patients for remote review, we estimated the effect of remote review on provider time and CGM TIR by comparing patients who did and did not receive remote review. We hypothesized that: 1) clinic patient capacity would increase as a result of changes in time spent on review and patient contacts, and that 2) remote review and individual contacts would improve patient’s CGM glucose TIR.

## METHODS

### Study design

This study consisted of two stages: a retrospective cohort review as part of a non-randomized quality improvement effort in an academic, pediatric T1D clinic; this was followed by a prospective case-control evaluation of the clinical workflow and patient glucose management. The Stanford University Institutional Review Board approved this work. Patients and families gave informed consent (or assent) prior to participating.

### Settings and participants

The work was conducted at a pediatric T1D clinic at a large pediatric academic medical center. Participants included all new T1D patients that chose to enroll in the remote review program. Participants in this program were provided with 1 month of CGM supplies and insurance approval was obtained for ongoing coverage as part of a research protocol. Participants who did not have their own iOS devices were provided with an iPod Touch to facilitate data sharing. Since this analysis is part of an ongoing pilot study, additional participants were added weekly.^18^

Before the work described here, the clinic was performing manual weekly CGM review of patients enrolled in the study.^1^ Our study and datasets are split into four periods: Period 0 is the historical period used to simulate, evaluate and define new review criteria; Period 1 immediately precedes the introduction of our new review criteria and serves as the baseline we use to measure the effects of our changes; Period 2 begins after our new review criteria are deployed in the clinic; and Period 3 is the measurement period after we deploy an interactive dashboard to simplify the review workflow.

### Clinical metrics, targets, and the patient priority ranking

The new criteria to prioritize patients for data review and a potential telehealth visit were generated based on a modified version of consensus guideline targets.^15,17^ Standard CGM metrics were used in this study: percent-worn (PW) is the proportion of time with recorded glucose readings, time-in-range (TIR) is defined as glucose readings 70-180 mg/dL, clinically significant hypoglycemia (csHyp) is defined as glucose readings below 54 mg/dL, and hypoglycemic (Hyp) is defined as the glucose readings below 70 mg/dL. A patient must have a PW ≥ 75% to be eligible for remote review during each study week. For the eligible patients, we defined three potential “flags” that could be raised for each patient each week: (1) TIR < 65%, (2) csHyp > 1%, and (3) Hyp > 4%. Patients with insufficient readings were contacted to understand the reason for missing measurements (e.g. out of supplies, technical issues, taking CGM break).

Patients were ranked (i.e., prioritized for review) in descending order of number of flags. Within groups of patients that have the same number of flags, patients were ranked in ascending order of TIR. This combined ranking is what we subsequently deployed in our dashboard in Period 2 to replace the existing review criteria in Period 1, which involved reviewing all patients with TIR < 70%, csHyp > 1%, or Hyp > 4%.

### Creation and evaluation of criteria for review based on retrospective data

In the first stage of the study, we developed criteria for review using a historical dataset from patients in our clinic. During this historical period, the data of patients with and without any of the flags described above were reviewed remotely by a Certified Diabetes Educator (CDE). The historical dataset included weekly data on which patients had their CGM data reviewed by a CDE and which patients had been contacted as a result of this review. We estimated the priority rank of each patient in each of the previous weeks they had passively uploaded CGM data.

CDEs were presented with a choice for how many patients to review weekly, K, (ordered by our ranking function) and the sensitivity and specificity corresponding to each choice. We generated a receiver operating characteristic (ROC) curve that graphs the positive predictive value (PPV; precision) and sensitivity for each possible value of K. PPV is defined as the proportion of patients meeting the review criteria that were contacted in the historical data. Sensitivity is defined as the proportion of patients contacted historically that met the evaluated review criteria.

As K increases, more patients are reviewed and the CDEs are more likely to review all the patients they would contact if they reviewed them. But there is also a cost associated with increasing K: CDE resources are limited, and so there is a limit to the clinic population they can monitor. Small values of K correspond to the potential for more patients to be monitored given a fixed amount of CDE time.

We simulated an additional option to review all patients who had flags over the past two consecutive weeks along with the top K patients according to the ranking function, in order to ensure that patients with flags in two consecutive weeks were reviewed despite ranking below the top K.

The hypothetical number of patients reviewed weekly along with sensitivity estimates of each candidate criteria were presented to the clinical staff along with a comparison against the existing review criteria in Period 1. The new criteria they chose was implemented in a dashboard used by the clinic to prioritize which patients to review each week

### Deployment in an academic, pediatric T1D clinic

The new review criteria were first deployed in Period 2 in the R Shiny dashboard the clinic was already using during Period 1 to determine which patients’ CGM data the CDEs should review each week (Figure 1). The change did not require modifying the existing workflow of the CDEs. The system that delivers patient CGM data to the clinic via Dexcom Clarity, as well as the clinic’s R Shiny dashboard, is described in Scheinker et al.^1^

**Figure 1:**
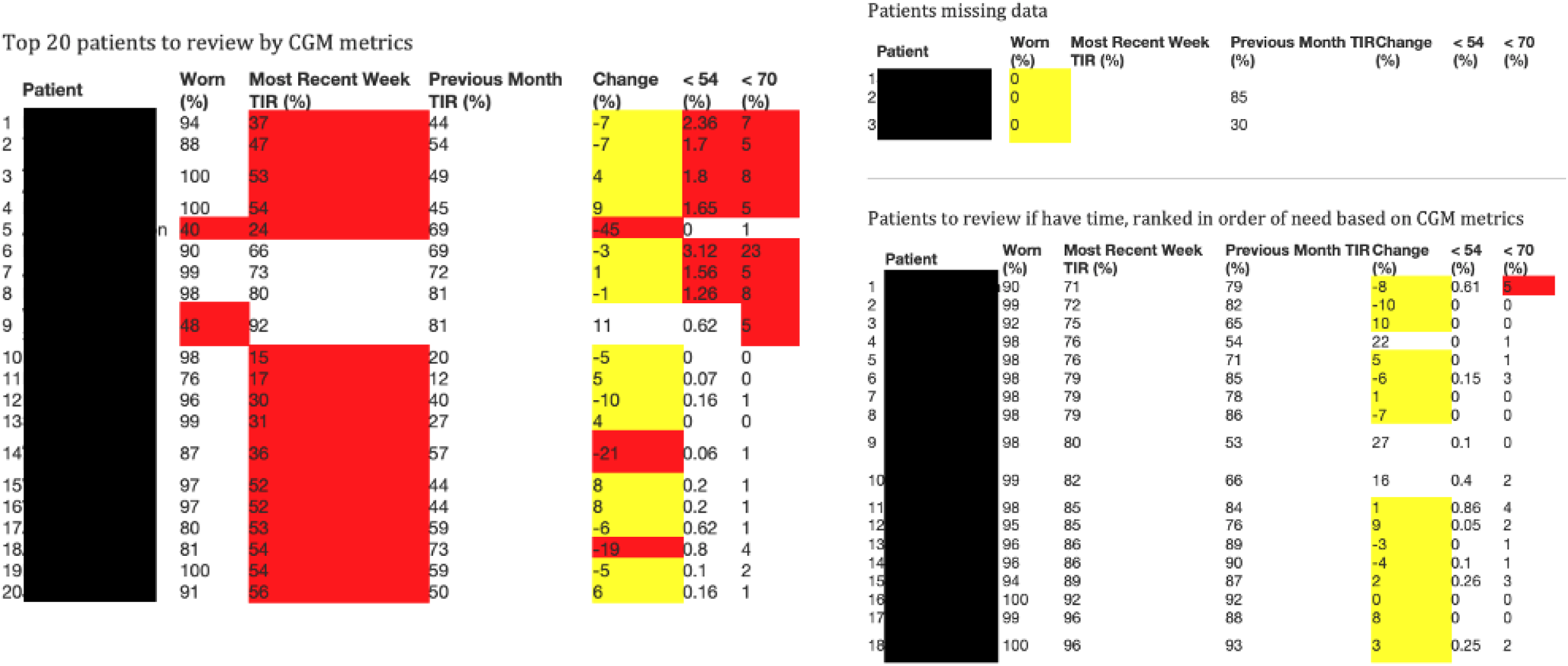
R Shiny Dashboard. Patients are ranked according to our new criteria. Individual patient’s data has to be reviewed separately in a web browser. The later interactive dashboard (Figures 5 and 6) removed the need to open a separate view to review individual patients.

The criteria were later deployed in an interactive Tableau dashboard in Period 3 (Figures 2 and 3). Unlike the R Shiny dashboard, the Tableau dashboard also showed each patient’s glucose data, including the Ambulatory Glucose Profile (AGP) view that quickly summarizes their data.^19^

**Figure 2:**
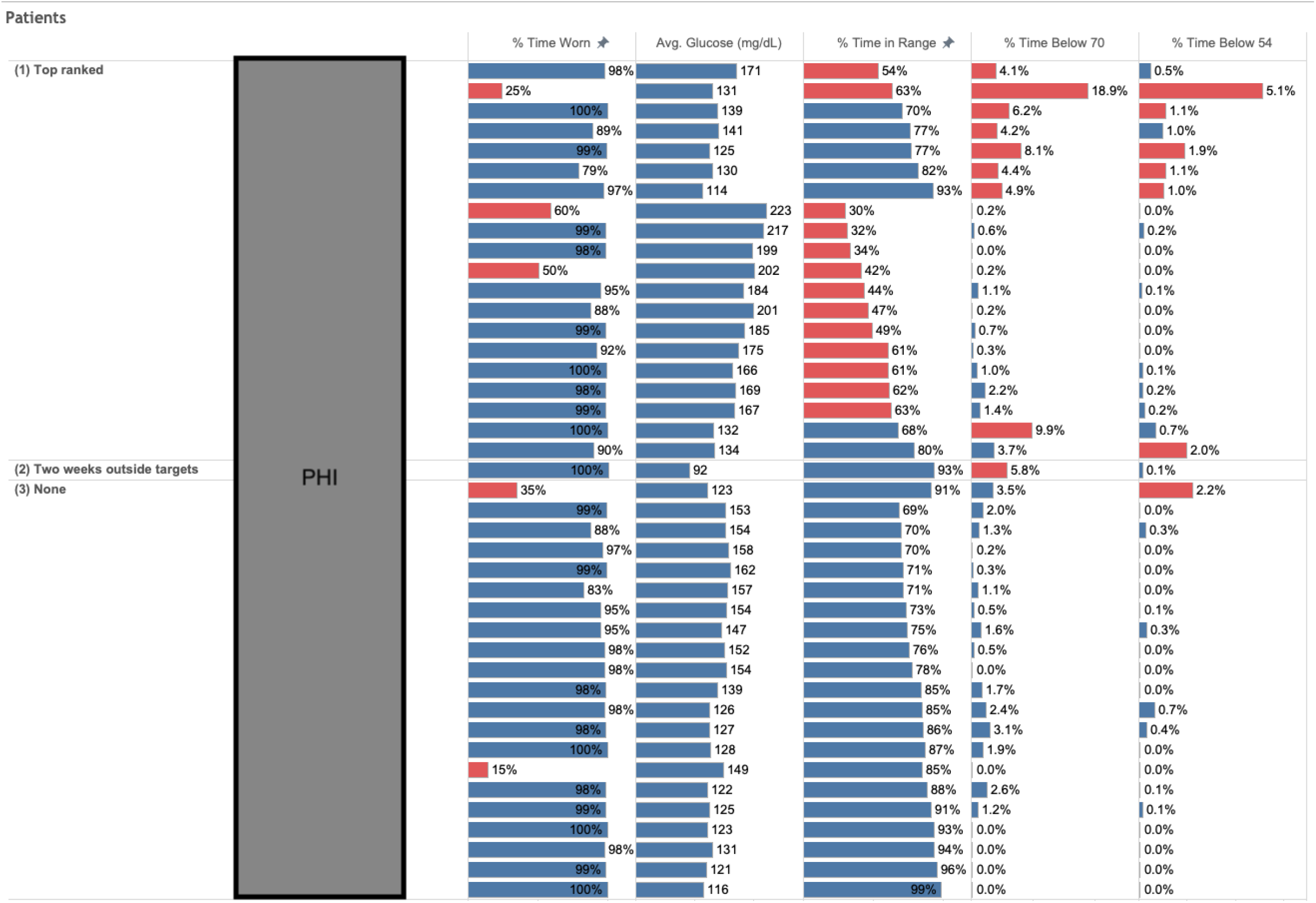
Interactive Tableau Dashboard (Population View) Patients are ranked according to our new clinical criteria. Once a CDE clicks on a patient, metrics and graphs below the table update to show a summary of that patient’s CGM data (see Figure 6).

**Figure 3:**
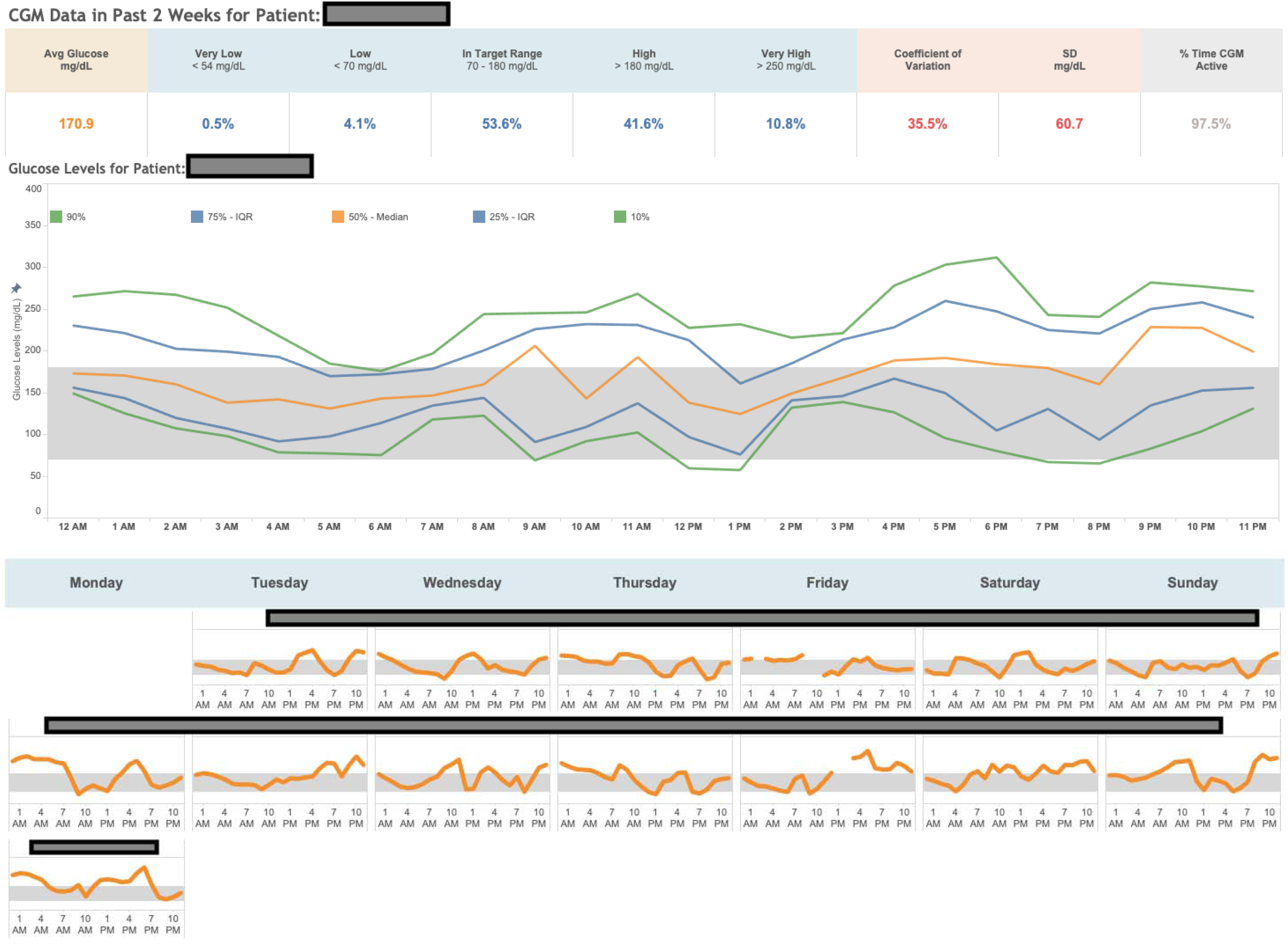
Interactive Tableau Dashboard (Patient View) This view updates to show data for the patient selected in the population view (Figure 2).

Showing these data was intended to eliminate the need for the CDEs to log into a separate web portal (Dexcom Clarity) to review patients’ data and to reduce the time needed to review each patient.

The time spent on reviewing patients’ CGM data and on contacting patients was tracked by the CDE performing the data review. The steps involved in this process were identifying a patient with a flag, opening Dexcom Clarity (for the R Shiny tool, but not the Tableau tool), reviewing the patient’s data; then if the CDE believed the patient needed a contact, they would open up the EHR, send a secure message, and potentially update insulin dosages in the chart.

After the new criteria took effect in Period 2, we continued recording the number of patients eligible for review each week, the number flagged for review, and the number contacted each week. Before and after the interactive dashboard was deployed in Period 3, CDEs reported the average time they spent reviewing a patient’s CGM data and the average time they spent contacting each patient after data review (sending messages and updating charts). We estimated the total time spent on review and contacts each week by multiplying the times reported per patient by the CDEs by the number of patients reviewed and contacted each week.

### Outcomes

The primary outcome for this study was the change to clinic patient capacity as a result of changes in time spent on review and patient contacts. This operational metric was estimated as the total amount of CDE time available weekly divided by the time necessary to review CGM data and make contacts per patient eligible for remote review in a given week.

A secondary outcome was the effect of remote review and individual contacts on glucose management. This was measured in three ways: (1) the weekly TIR at twelve months since first diagnosis of diabetes compared to a control cohort in the same clinic, (2) the difference in weekly TIR between patients with and without contacts in the preceding week, controlling for the TIR in the week preceding contact, and (3) the difference in the odds that a patient’s TIR improved by 5pp or more between patients with and without contacts in the preceding week, controlling for the TIR in the week preceding contact.

### Estimating the effect of remote review and contacts on glucose management

We compared the mean TIR in each month since diabetes onset among the clinic cohort with remote review to a cohort in the same clinic without remote review. Patients were included if they started CGM within a month after diagnosis, submitted CGM measurements a year after diagnosis, and had consented to the relevant IRB protocol. The primary reason some patients were ineligible for remote review was that their diabetes diagnoses happened before the clinic had received IRB approval for remote monitoring.

We performed regression modeling of the TIR during a patient-week to estimate the mean difference in TIR associated with a remote contact in the previous week while controlling for the patient’s TIR two weeks earlier. We fit two linear regression models: one with patient-level random effects and one with patient-level fixed effects. Beyond the ranking, CDEs’ choices of which patients to contact depends on the patients’ state; to further control for this effect, we fit a propensity model of contact and estimated the effects of contacts using a causal forest model (Supplement).^20^

We performed logistic regression modeling of the binary outcome that the change in TIR for a patient is 5pp or greater between the week preceding a potential contact and the week following a potential contact, estimating the relative odds ratio of a ≥5pp change when there was a contact in the middle week relative to when there was no contact, controlling for the TIR of the week preceding contact and patient-level random/fixed effects (Supplement).

## RESULTS

### Analysis of historical data and definition of new review criteria

The population analyzed grew from 53 patients in Period 0 (historical data used to define new review criteria) to 225 patients in Period 3 (after deployment of the interactive dashboard for patient review). Cohort characteristics across periods are summarized in Table 1.

**Table 1:**
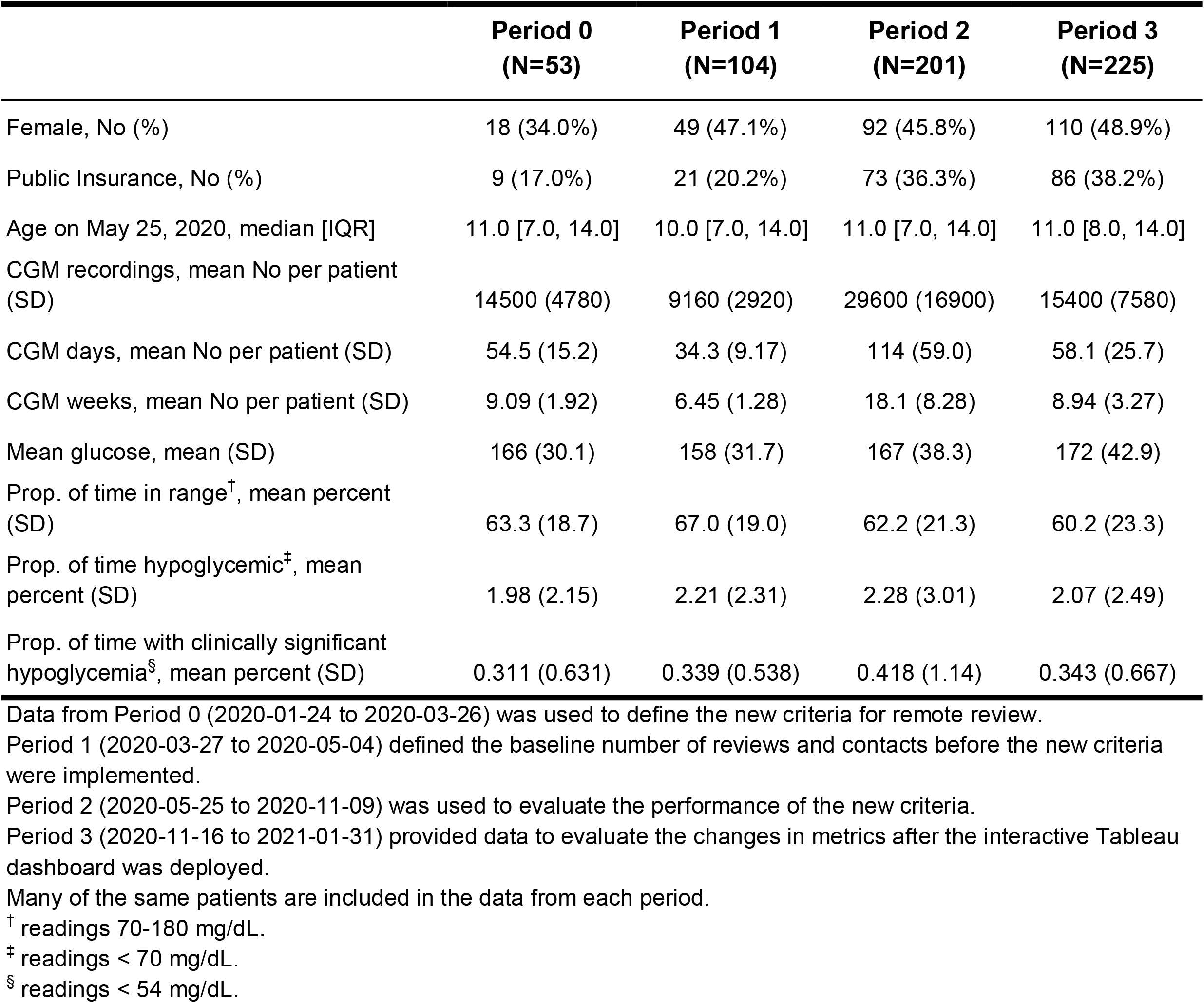
Population Summary Statistics by Time Period.

When using the criteria from Period 1, which mandated a manual review for each patient with a flag, CDEs reviewed data from 26 patients weekly on average. This number decreased to an estimated 22 patients with the new criteria: 20 patients selected by the ranking function and 2 patients with flags over the past two consecutive weeks (Figure 4).

**Figure 4:**
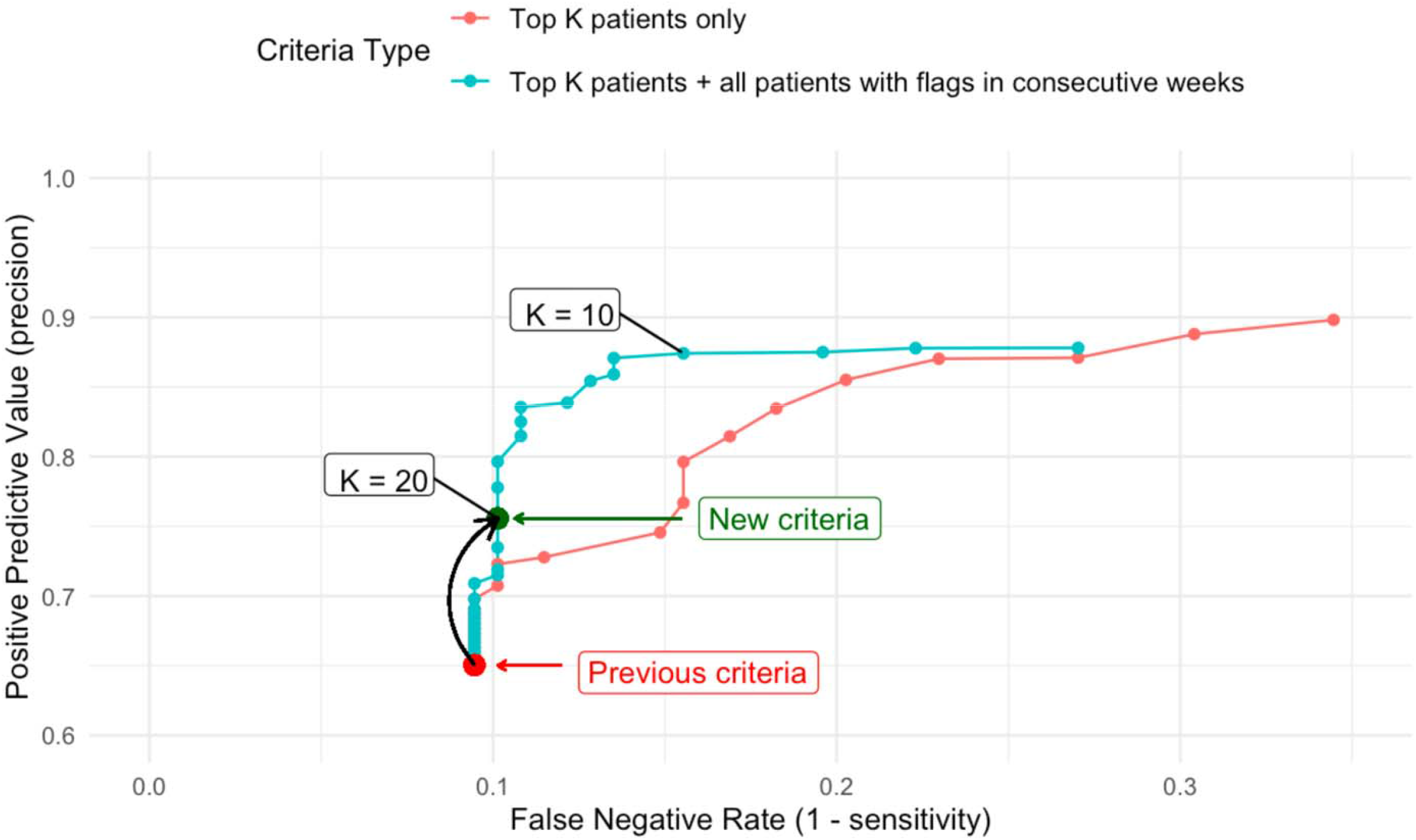
Receiver Operating Characteristic Curve Showing the Estimated Improvement in Positive Predictive Value from a Change in Review Criteria, Based on Historical Data. Our change moved our historical performance from the red point (previous criteria) to the green point (new criteria), increasing the historical positive predictive value from 0.65 to 0.76 while only reducing historical sensitivity from 0.91 to 0.90. This figure was generated using data from Period 0 in Table 1.

### Deployment in an academic, pediatric T1D clinic

After adopting the new criteria described above within the clinic’s R Shiny dashboard in Period 2 (Figure 1), the number of patients flagged each week dropped to 26 the following week. In the following months, the number of patients eligible for review increased slightly but the number of patients contacted remained stable (Figure 5).

**Figure 5:**
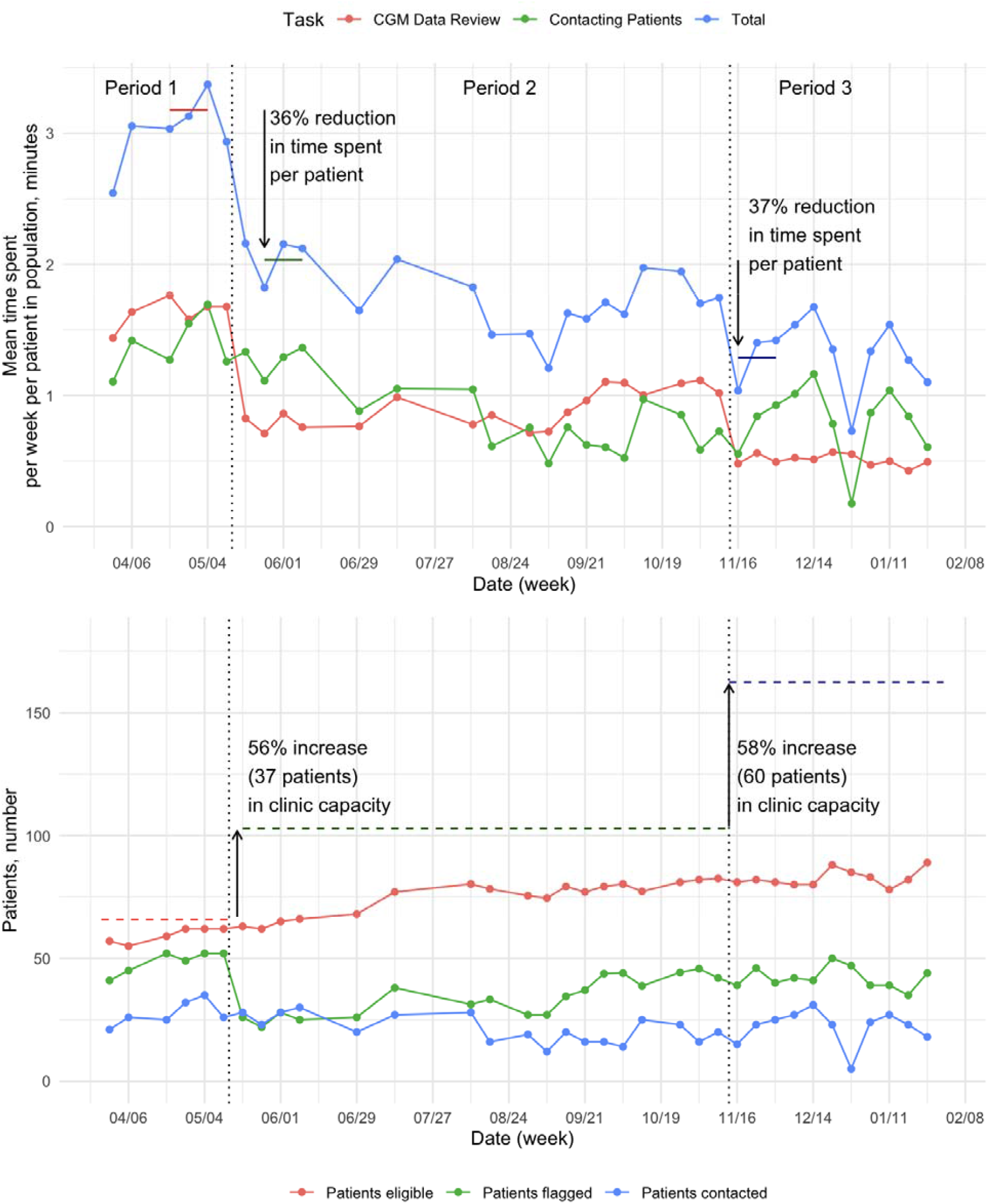
Time Saved, Increase in Clinic Capacity and Patients Eligible after Tool Changes. In both panels: the first vertical dashed line from the left shows the time of the review criteria change. The second line shows the time of the Tableau tool deployment. In the bottom panel: the clinic total time budget is fixed to the maximum time spent in a week before the criteria change (209 minutes). Clinic capacity is estimated by dividing the total time budget by the time spent per patient per week. The top panel shows the changes in the time spent per patient per week and the bottom panel translates those changes into changes in estimated clinic capacity.

Using the per-patient CDE-reported time estimates for CGM data review (2 minutes per patient reviewed), and EMR-based updates and messaging (3 minutes per patient contacted) with our tool, we estimated the time spent reviewing and messaging patients each week before and after the criteria change. The estimated time spent per patient eligible for review in the population dropped 36% from 3.2±0.20 to 2.0±0.21 minutes in the three weeks after the criteria change as compared to the three weeks before the change (Figure 5). This time reduction resulted from the lower number of weekly patient reviews with the revised criteria (Figure 5).

With an estimated fixed weekly 209 minutes available for reviewing CGM data and contacting patients, the measured 36% reduction in time spent per patient corresponds to a 56% increase in the estimated maximum clinic patient capacity (from 66 to 103 patients; Figure 2).

In November 2020, we deployed an interactive Tableau dashboard (Figures 2 and 3) to replace the clinic’s R Shiny dashboard (Figure 1). Based on self-reported time measurements from the clinic’s CDEs, the new dashboard reduced the mean time CDEs spent reviewing each indicated patient’s CGM data from 2 minutes to 1 minute. CDEs could review each patient’s CGM data on a single platform, rather than the previously required access to a separate webpage for each patient. As a result, the estimated time spent per patient in the population dropped 37% from 2.0 to 1.3 minutes and the estimated clinic patient capacity increased by an additional 58%.

When combined, these two changes reduced the estimated per patient time spent on review and contact by 60%, from 3.2±0.20 to 1.3±0.24 minutes and increased the estimated clinic capacity by 147% (97 patients).

### Estimating the effect of remote review and contacts on glucose management

The mean TIR at 12 months since diagnosis was 65.8% for patients eligible for remote review and 57.0% for those who were enrolled in the study prior to remote review being established, which represents an improvement of 8.8pp (CI = 0.6–16.9pp) (Figure 6)

**Figure 6:**
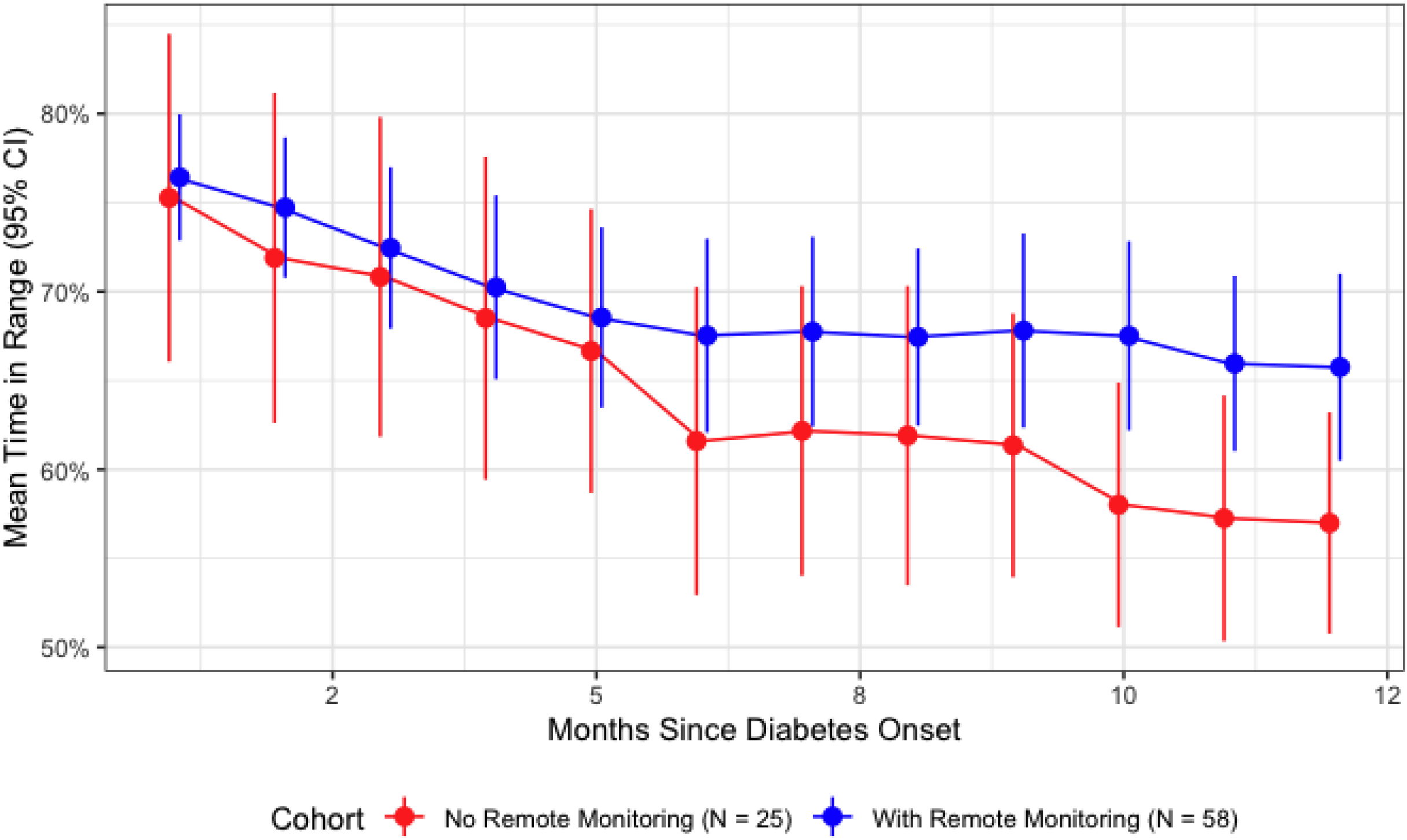
Mean Time in Range by Time Since Diabetes Onset, Comparing Cohorts with and without Remote Monitoring. The mean TIR in the remotely monitored cohort is 65.8% (95% CI: 60.5%, 70.9%) at 12 months since diabetes onset versus 57.0% (95% CI: 50.9%, 63.1%) in the cohort without remote monitoring. The 95% CI of the difference in the means at 12 months from a two-sample t-test is [0.6%, 16.9%] and the p-value is 0.0357. Note that remote monitoring was not assigned randomly so we cannot make a causal claim here. Summary statistics (remote monitoring vs no remote monitoring cohort): 27/58 vs 13/25 female, 14/58 vs 0/25 public insurance, 10.1 (SD: 4.7) vs 9.6 (SD: 5.1) years mean age on May 25, 2020.

Contact with a care provider was associated with an increase in the TIR the following week of 1.6pp (P = 0.02) in a model with patient random effects and an increase of 2.3pp (P < 0.001) in a model with patient fixed effects. These effect estimates were consistent with the results of a causal forest model (details in Supplement).

Contact with a care provider was associated with a greater chance of a TIR improvement of 5pp or more, with an odds ratio (OR) of 1.29 (CI = 1.02–1.64; P = 0.03) in a model with patient random effects and OR of 1.34 (CI = 1.05–1.71; P = 0.02) in a model with patient fixed effects (details in Supplement).

## DISCUSSION

In a pediatric T1D clinic at an academic medical center, the use of an algorithmic approach to identifying patients most likely to benefit from an asynchronous telemedicine contact was associated with a 60% reduction in the per-patient time required for review and contact and 29-34% increased odds of improving time-in-range by 5pp during the week following contact. The algorithm was based on clinician-developed review criteria. Its use decreased the proportion of patients in a clinic receiving remote provider CGM review each week without reducing the number of clinically indicated patient contacts. The improved efficiency corresponded to an estimated 147% increase in clinic capacity, from 66 patients to 163 patients, without requiring additional resources, and while improving patient glucose control.

Algorithm-enabled patient prioritization safely frees up provider time relative to reviewing all patients using CGM, which is not currently done due to it not being feasible. Consider a patient with TIR just below the consensus target one week and above target in the following week. The patient would not be reviewed if there are many other patients further outside of glucose targets or experiencing poor glucose control over longer time periods. These improvements enable an expansion of the patient population eligible for remote CGM review. Further, our results suggest a strong association between remote review and contact, and subsequent improvement in patient glucose control. Use of this algorithm facilitates a precision medicine approach translated to a clinic population in which patients receive clinician attention based on CGM data.

The tool described has been made available as open-source software.^21^ Our team will provide clinical and operational support for adoption by other clinics. The guideline-based inputs to the ranking function are fully interpretable and in-line with the latest consensus guidelines. Clinics already monitoring patients using CGMs may calibrate the tool to fit their preferred sensitivity/specificity trade-off. A clinic without a remote monitoring program may use the default settings of the tool. Earlier work in the development of this tool suggests that the types of alerts used may be calibrated based on the glucose management of the population cared for by the clinic.^1^

For identifying patients that had previously been contacted by CDEs after they reviewed the patients’ CGM data, in our clinic we found that a target of 65% TIR optimized specificity.^1^ This decision was supported by our clinical collaborators as an acceptable way to more efficiently target patients in need of intervention. Our target use of a 65% TIR target in the algorithm, which differs from the consensus guideline 70% TIR target, is one example of how the algorithm may be tailored to specific clinic populations and practices. Future directions include adjustment of this target to improve TIR and maintain efficiency.

We employed widely used tools (Python, Dexcom Clarity, R and Tableau) to produce these results. To our knowledge, existing CGM review tools either focus only on individual patients, are not open source, and/or show clinic population statistics only for patients wearing the same type of sensor.^22^ Our tool is open source and can work with CGM data from any source that supports raw data exports. These algorithms require minimal input and can be run on an ordinary laptop. Barriers to the implementation at scale of algorithm-enabled improvements to T1D care will more likely be associated with ancillary difficulties (secure data access, data provision by device manufacturers, and agreement with local regulations), than with the technical challenges of using it. Policymakers, patients, physicians, and other stakeholders should engage with the issue of data sharing to ensure that the failures that plague EMR interoperability and data sharing do not befall CGM data.^23^ In the US, the 21^st^ Century Cures Act provides a framework to facilitate patient, family, and provider access to medical records and electronic health information.^24^

Our work facilitates the expansion of remote CGM review to larger patient populations and potentially to smaller primary care clinics. While increases in clinic capacity present an opportunity to expand access among disadvantaged populations, a thoughtful implementation is paramount to avoid the risk of intervention-generated inequalities.^25^ The ADA recommends CGM for all youth with T1D, but currently access to CGM is limited for youth on public insurance, increasing disparities. Access to CGM is a first step, and needs to be combined with diabetes education, as well as collaboration with the diabetes care team to assist with insulin dose adjustments as needed to achieve tight glucose control. Numerous steps are necessary to ensure that underserved populations reap the benefits associated with care enabled by new technologies.^7,26^

Further research is required to evaluate how our methods can be best applied in a clinic with multiple providers reviewing CGM data. A clinic-level prioritization tool may generate additional benefits when multiple providers are involved with patient care, rather than a split cohort setting like ours. Such a tool may also support and standardize the training and integration of new providers. Including insulin data in the tools used by CDEs could help inform treatment decisions. Better understanding of how individual patients respond to remote contacts and adding personalized CGM targets for patients may unlock further efficiencies in how provider resources are allocated within the clinic.

The limitations of our work include a small sample population size of mostly new onset patients, reducing our ability to detect small changes in patients’ glucose measurements or HbA1c that resulted from our work. Further, time allocation and capacity calculations were based on estimates from experienced CDEs. Prospectively collected objective measurements of time allocation would strengthen future studies. Note that our results on increased clinic capacity are fairly robust to even large errors in the CDE time estimates per patient, as they rely primarily on safe decreases in the number of patients requiring review each week and secondarily on marginal time savings associated with a simpler review workflow. Our estimation of the effect of provider contacts is limited by remote review not being randomly assigned to patients, which limits our ability to make causal claims. We also cannot conclude how our work would translate to primary care, adult T1D, or type 2 diabetes clinics.

In conclusion, an algorithm-enabled prioritization of T1D patients with CGM for asynchronous remote review reduced provider time spent per patient by over 60% and was associated with improved TIR. This demonstrates that a pediatric T1D clinic with fixed resources may be able to increase its capacity and TIR through standardized, population-level, algorithm-enabled CGM data review and telemedicine. Algorithmic improvements to clinic capacity may present an opportunity to expand access to remote CGM review, especially to underserved populations without local access to pediatric endocrinologists.

## Data Availability

Some of the summary data are available at request from the corresponding author. The raw patient data is protected by privacy laws and cannot be shared.

## Acknowledgements

We thank the staff of the T1D clinic in which the study was conducted. We also thank Josh Grossman and Saied Mehdian for their advice on this work.

## Supplement

### Estimating the effect of remote review and contacts on glucose management

Regression specification to estimate effect of contacts on TIR:

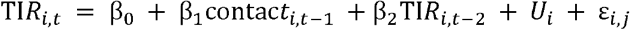

*TIR*_*i,t*_ is the proportion (0-1) of time patient i was in range in week t. *β*_0_ is the estimated intercept. *contact*_*i,t-*1_ is an indicator variable equal to 1 if patient i received a remote contact in week t-1 and equal to zero otherwise. *β*_1_is the coefficient of interest, representing the estimated effect of a remote contact in week t-1 on TIR in week t. *U*_*i*_ is the patient-level random or fixed effect depending on the estimated model. *ε* _*i,j*_ is the residual.

We fit two models using the R plm package, one with patient-level fixed effects and one with patient-level random effects.^1^ Random effects were fit using the default *swar* method.^2^ To also test a non-linear model, we fit a causal forest to estimate the average effect of remote contact on TIR. We used the R grf package to estimate the average treatment effect on the treated.^3^ This involved first trained a random forest regression model on the control variable *TIR*_*i,t*-2_ to estimate the propensity of contact in the previous week *contact*_*i,t*-1_. Then, the causal forest model predicts individual treatment effects based on the estimated propensities and the control *TIR*_*i,t*-2_. Finally, we estimate the average treatment effect across the treated (i.e. contacted) patient-weeks. The hyperparameters of both forest models were tuned using the grf package’s built-in automatic tuning capabilities.

The sample used to train the models contained 3,657 patient-weeks and 113 unique patients. Only patients that had been tracked for at least twelve weeks were included, and the sample was limited to observations with *TIR*_*i,t*-2_ < 0.7 to remove patients that already have good glucose control when eligible for contact. 525 of the observations had *contact*_*i,t*-1_ = 1.

The estimated effects of contacts on TIR is 1.6pp (SE: 0.7pp) in the linear model with patient random effect, 2.3pp (SE: 0.7pp) in the linear model with patient fixed effect, and 1.7pp (SE: 0.7pp) in the causal forest (Table A1). For the regression models, the estimated coefficients 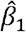, their standard errors and p-values are reported in Table S1. For the causal forest, the estimated average treatment effect on the treated and its standard error is reported.

Logistic regression specification to estimate effect of contacts on the odds that patient’s TIR increases by 5pp or more:

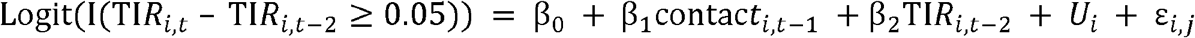

The variable definitions are identical to those described in the earlier regression specification. I (TIR_*i,t*_ – TIR_*i,t*-2_ ≥ 0.05) is an indicator variable equal to one when the change in TIR of patient i increases by 5pp or more from week t-2 to week t. exp (β_1_) is the parameter of interest, which estimates the odds ratio (OR) of a TIR improvement of 5pp or more when there is a contact in the middle week relative to when there is no contact, controlling for the TIR of the first week. The model was fit with same sample as before, and a model was fit separately with patient-level random effects and with patient-level fixed effects. The estimated ORs, their 95% confidence intervals and p-values are reported in Table S2.

**Table S1:**
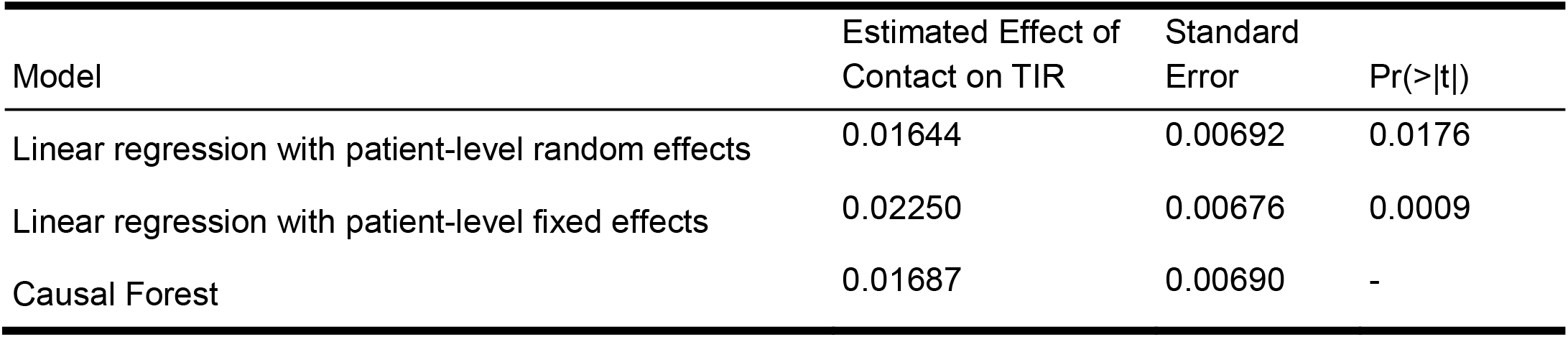
Estimated effects of remote contacts on patients’ time in range (TIR)

**Table S2:** Estimated effects of remote contacts on the odds that TIR improves by ≥ 5pp.

| Model | Estimated effect of Contact (Odds ratio) | 95% CI | Pr(>\|z\|) |
| --- | --- | --- | --- |
| Logistic regression with patient-level random effects | 1.292 | 1.020–1.637 | 0.0329 |
| Logistic regression with patient-level fixed effects | 1.341 | 1.051–1.712 | 0.0182 |

## Footnotes

1 Croissant Y, Millo G (2008). “Panel Data Econometrics in R: The plm Package.” Journal of Statistical Software, 27(2), 1–43. doi: 10.18637/jss.v027.i02.

2 Swamy and Arora 1972

3 Wager S, Athey S. Estimation and inference of heterogeneous treatment effects using random forests. Journal of the American Statistical Association. 2018 Jul 3;113(523):1228-42.

